# Hydroxychloroquine and azithromycin: As a double edge sword for COVID-19?

**DOI:** 10.1101/2021.01.16.21249941

**Authors:** Seyed Parsa Eftekhar, Sohrab Kazemi, Mohammad Barary, Mostafa Javanian, Soheil Ebrahimpour, Naghmeh Ziaei

**Affiliations:** Student Research Committee, Health Research Center, Babol University of Medical Sciences, Babol, Iran; Cellular and Molecular Biology Research Center, Health Research Center, Babol University of Medical Sciences, Babol, Iran; Infectious Diseases and Tropical Medicine Research Center, Health Research Institute, Babol University of Medical Sciences, Babol, I.R. Iran; Department of Cardiology, Babol University of Medical Sciences, Babol, Iran

**Author notes:** **Corresponding Author**: Naghmeh Ziaei, Department of Cardiology, Babol University of Medical Sciences, Babol, Iran, E- mail.

**Keywords:** Hydroxychloroquine, Azithromycin, QTc interval, Torsade de pointes, COVID-19, novel coronavirus

## Abstract

**Background:** Hydroxychloroquine with or without azithromycin was one of the common therapies at the beginning of the COVID-19 pandemic. They can prolong QT interval, cause Torsade de pointes, and lead to sudden cardiac death. We aimed to assess QT interval prolongation and its risk factors in patients who received hydroxychloroquine with or without azithromycin.

**Methods:** This was a retrospective cohort study. 172 patients with COVID-19 included, hospitalized at hospitals of Babol University of Medical Sciences between March 5, 2020, and April 3, 2020. Patients were divided into two groups: hydroxychloroquine alone and hydroxychloroquine with azithromycin. Electrocardiograms were used for outcome assessment.

**Results:** 83.1% of patients received hydroxychloroquine plus azithromycin vs 16.9% of patients who received only hydroxychloroquine. The mean age of patients was 59.2 ± 15.4. The mean of post-treatment QTc interval in the monotherapy group was shorter than the mean of post-treatment QTc interval in the combination therapy group but it had no significant statistical difference (462.5 ± 43.1 milliseconds vs 464.3 ± 59.1 milliseconds; *P* = 0.488). Generally, 22.1% of patients had a prolonged QTc interval after treatment. Male gender, or baseline QTc ≥ 450 milliseconds, or high-risk Tisdale score increased the likelihood of prolonged QTc interval. Due to QTc prolongation, 14 patients did not continue therapy after 4 days.

**Conclusion:** Hospitalized patients treated with hydroxychloroquine with or without azithromycin, had no significant difference in prolongation of QT interval and outcome. But the number of patients with prolonged QT intervals in this study emphasizes careful cardiac monitoring during therapy; especially in high-risk patients.

## 1 Introduction

COVID-19 spread fast and infected millions of people over the world until now from its beginning in Wuhan, China, and causes high mortality over the world, especially in patients with underlying diseases such as diabetes and congestive heart failure[1, 2]. Since enough clinical trials were unavailable at the early stages of this pandemic, protocols suggested different treatments based on repurposing of medications, clinical trials, in vitro studies, and experiences from past coronaviruses like SARS and MERS; However, definite treatment and vaccine need more time and work[3-6]. Remdesivir, hydroxychloroquine sulfate, chloroquine phosphate, and many other drugs showed antiviral efficiency in vitro. Even Yao and colleagues suggested hydroxychloroquine is more potent than chloroquine[7, 8]. Shah et al. suggested prophylactic roles for chloroquine and hydroxychloroquine[9]. But clinical trials challenged hydroxychloroquine efficiency and NIH (National Institutes of Health) denied hydroxychloroquine and chloroquine consumption except in clinical trials[10-12].

Hydroxychloroquine sulfate –which is used for malaria and autoimmune diseases like systemic lupus erythematosus and rheumatoid arthritis– with or without azithromycin was one of the common therapies at the beginning of the pandemic: Studies in vitro report hydroxychloroquine could inhibit virus fusion and entry to cell and confronts against the virus[7, 8]. The immunomodulatory effect of hydroxychloroquine is another reason[13, 14]. But a notable concern raised with this treatment.

Hydroxychloroquine sulfate and macrolides such as azithromycin affect normal electrophysiologic function of the heart and concurrent use, cause synergistic effect, and may lead to prolonged QT interval, torsades de pointes, and even death[15-18]. Due to the widespread and arbitrary use of these drugs, risk-assessment is necessary. However, some studies proceeded on hydroxychloroquine and macrolides effects on QTc interval, still a question has remained unanswered: does short-term consumption of hydroxychloroquine and azithromycin cause unwanted cardiac effects? We aimed to conduct a retrospective evaluation at hospitals of Babol University of Medical Sciences to assess different parameters of electrocardiogram such as QT interval duration and risk factors of QT interval prolongation to find out de-novo arrhythmias along with possible electrophysiologic adverse events among hospitalized adult patients with COVID-19.

## 2 Methods and Materials

### 2.1 Data sources

This was a retrospective descriptive survey among adult patients with COVID-19, hospitalized at hospitals of Babol University of Medical Sciences. All data and electrocardiograms were extracted from the patients’ files. Research was approved by the ethics committee of Babol University of Medical Sciences.

### 2.2 Patients

We included hospitalized patients between March 5, 2020, and April 3, 2020, and this period was chosen for enough patients with determined discharge status (alive or dead). Inclusion criteria were at least one positive real -time reverse transcriptase polymerase chain reaction test (RT-PCR) of nasopharyngeal samples and age over 18 years old. Exclusion criteria were age below 18 years old, severe metabolic disease, who were unable to receive oral drugs like lactation and pregnancy, and patients who were dependent on continuous renal replacement therapy like hemodialysis.

### 2.3 Drug regimen

All patients received oral hydroxychloroquine sulfate 600 mg daily (200 mg three times daily for 10 days); and azithromycin initiated for 141 patients (500 mg on day one and 250 mg for the next four days). We divided patients based on the pharmaceutical diet into two groups: (1) hydroxychloroquine alone and (2) hydroxychloroquine with azithromycin.

### 2.4 Outcome assessment

Electrocardiograms were used for outcome assessment of the study. Electrocardiograms were described manually by a cardiologist. Baseline electrocardiograms were recorded at the time of admission. Several electrocardiograms were recorded after treatment, but we used electrocardiograms recorded 3 hours after the second dose of treatment (as post-treatment electrocardiograms) (Based on QTc monitoring guidance by Massachusetts General Hospital)[19]. QTc interval was calculated by the Bazett formula. The Tisdale Risk Score –which predicts QT interval prolongation risk in hospitalized patients– was used for risk assessment of developing prolonged QT interval.

### 2.5 Statistical analysis

Proportions were used to represent nominal variables. Means and standard deviations were used to represent continuous variables. χ2 or Fisher exact test was used for comparing categorical variables and odds ratio (ORs) and 95% CI were used for describing them. Continuous variables were compared using the Mann-Whitney U test, and *P* value < 0.05 was considered as a statistical significance threshold. The logistic regression model was used for evaluating QTc interval prolongation risk (ΔQTc ≥ 60 milliseconds or post-treatment QTc interval > 500 milliseconds). SPSS version 20 was used for statistical analysis.

## 3 Results

143 (83.1%) patients received hydroxychloroquine plus azithromycin vs 29 (16.9%) patients who received only hydroxychloroquine. The mean age of patients was 59.2 ± 15.4. About half of the patients were female. The most common comorbidities were diabetes mellitus, hypertension, and congestive heart failure. More than half of the patients had no history of cardiovascular diseases. The mean of Tisdale score was 8 ± 2.1 and 12.2% had high-risk scores (score ≥ 11) (Table 1).

**Table 1.** COVID-19 patients’ characteristics at the time of administration. Abbreviations: SD, Standard Deviation

| Characteristic | Total (n=172), (%) | Hydroxychloroquine (n=29), (16.9%) | Hydroxychloroquine and azithromycin (n=143), (83.1%) | P value |
| --- | --- | --- | --- | --- |
| Age, mean ± SD, year | 59.2 ± 15.4 | 59.1 ± 17.9 | 59.8 ± 14.8 | 0.577 |
| Female | 85 (49.4) | 16 (55.1) | 69 (48.2) | 0.497 |
| Intensive care at time of testing | 17 (9.8) | 3 (10.3) | 14 (9.7) | 0.927 |
| Tisdale score at treatment initiation, mean ± SD | 8 ± 2.1 | 5.8 ± 2.3 | 8.4 ± 1.8 | < 0.001 |
| Acute heart failure | 28 (16.2) | 6 (20.6) | 22 (15.3) | 0.480 |
| Maximum temperature at the time of administration, mean ± SD, C° | 37.1 ± 0.5 | 37.1 ± 0.4 | 37.2 ± 0.5 | 0.192 |
| **Baseline laboratory values, mean ± SD** |  |  |  |  |
| Serum potassium, mEq/L, mean ± SD | 4.2 ± 0.3 | 4.3 ± 0.3 | 4.1 ± 0.3 | 0.062 |
| Serum creatinine, mg/dL, mean ± SD | 1.2 ± 1.1 | 1.1 ± 0.3 | 1.2 ± 1.2 | 0.810 |
| White blood cell count, cells/μL, mean ± SD | 7140.1 ± 3501.4 | 7848.6 ± 4880.4 | 6995.2 ± 3128.1 | 0.627 |
| Absolute lymphocyte count, count/μL, mean ± SD | 22.78 ± 9.6 | 23.1 ± 12.3 | 22.71 ± 9.1 | 0.325 |
| C-reactive protein, mg/dL, mean ± SD | 8.8 ± 5.9 | 9.6 ± 7.4 | 8.6 ± 5.6 | 0.708 |
| Creatine phosphokinase, IU/L, mean ± SD | 232.4 ± 395.5 | 146.4 ± 55.9 | 272.5 ± 473.7 | 0.940 |
| Lactate dehydrogenase, IU/L, mean ± SD | 642.9 ± 358.7 | 507.2 ± 219.3 | 679.1 ± 380.3 | 0.003 |
| **Preexisting conditions** |  |  |  |  |
| Hypertension | 53 (30.8) | 10 (34.4) | 43 (30.1) | 0.639 |
| Congestive heart failure | 48 (27.9) | 4 (13.7) | 44 (30.7) | 0.063 |
| Coronary artery disease | 11 (6.3) | 0 (0) | 11 (7.6) | 0.123 |
| Atrial fibrillation | 2 (1.2) | 1 (3.4) | 1 (0.7) | 0.208 |
| Diabetes mellitus | 60 (34.8) | 7 (24.1) | 53 (37.1) | 0.183 |
| Chronic obstructive pulmonary disease | 4 (2.3) | 0 (0) | 4 (2.7) | 0.362 |
| Malignancy | 7 (4.1) | 2 (6.8) | 5 (3.4) | 0.398 |
| Chronic kidney disease | 10 (5.8) | 1 (3.4) | 9 (6.2) | 0.550 |
| **Medications** |  |  |  |  |
| Loop diuretics | 41 (23.8) | 8 (27.5) | 33 (23.1) | 0.603 |

The mean of baseline QTc interval –before initiating the treatment– in the monotherapy group was more than the mean of baseline QTc interval in the combination therapy group but it was not statistically significant (median ± SD, 459.9 ± 38.3 milliseconds vs 447.1 ± 44.1 milliseconds; *P* = 0.198). The mean of post-treatment QTc interval in the monotherapy group was shorter than the mean of post-treatment QTc interval in the combination therapy group but it had no significant statistical difference (median ± SD, 462.5 ± 43.1 milliseconds vs 464.3 ± 59.1 milliseconds; *P* = 0.488). (Table 2) (Figure 2).

**Table 2.** Baseline and post-treatment QTc interval. Abbreviations: SD, Standard deviation; QTc, Corrected QT interval; ΔQTc, Change in corrected QT interval; ms, millisecond.

| Characteristic | Total (n=172), (%) | Hydroxychloroquine (n=29), (%) | Hydroxychloroquine and azithromycin (n=143), (%) | P value |
| --- | --- | --- | --- | --- |
| Baseline QTc, mean ± SD, ms | 449.1 ± 43.3 | 459.9 ± 38.3 | 447.1 ± 44.1 | 0.198 |
| Post-treatment QTc, mean ± SD, ms | 464.1 ± 56.6 | 462.5 ± 43.1 | 464.3 ± 59.1 | 0.488 |
| ΔQTc, mean ± SD, ms | 15 ± 50.3 | 2.6 ± 29.1 | 17.2 ± 53.4 | 0.463 |
| Prolonged QTc | 38 (22.1) | 3 (10.3) | 35 (24.5) | 0.094 |

**Figure 1.**
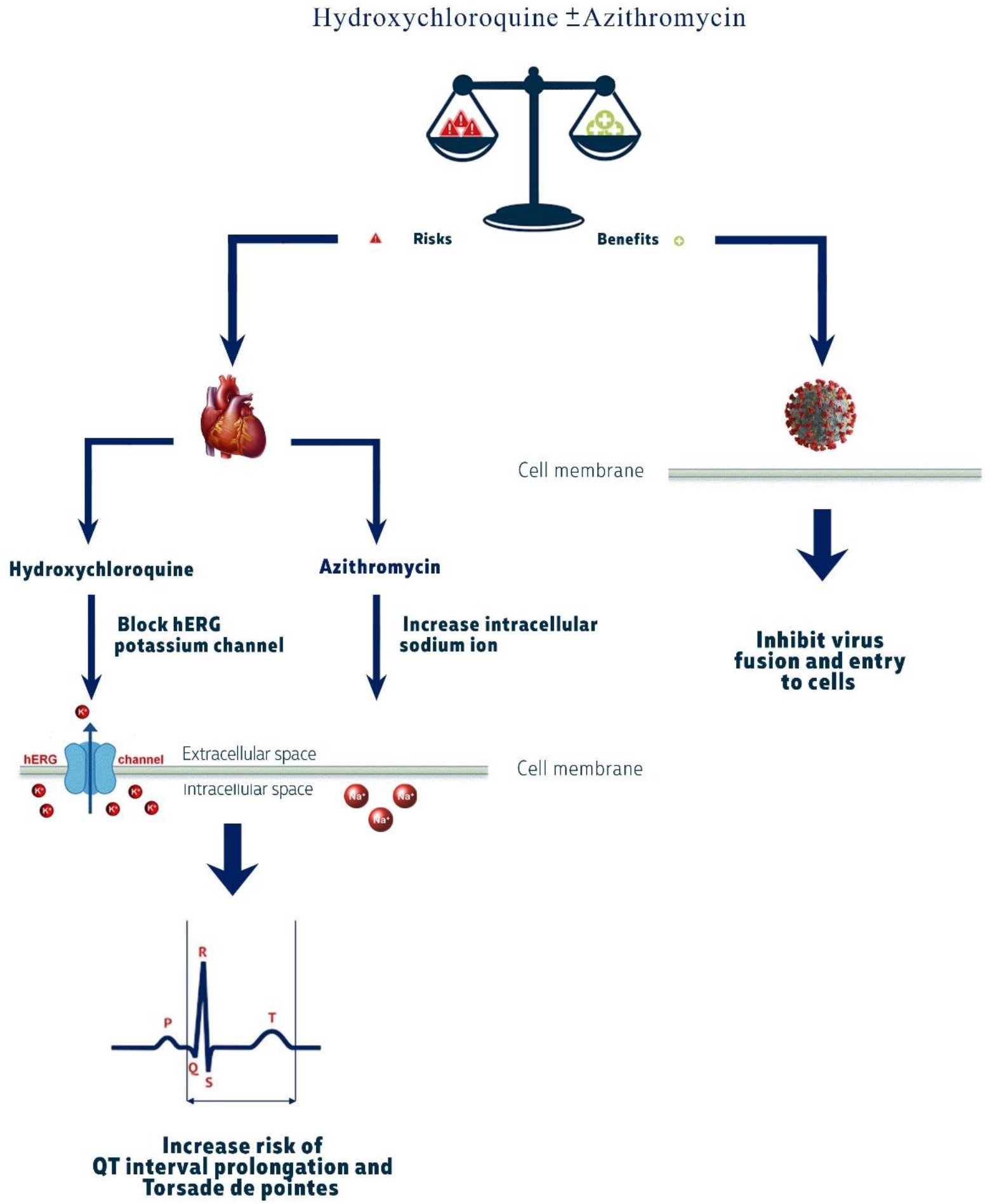
Hydroxychloroquine and azithromycin: As a double edge sword for COVID-19.

**Figure 2.**
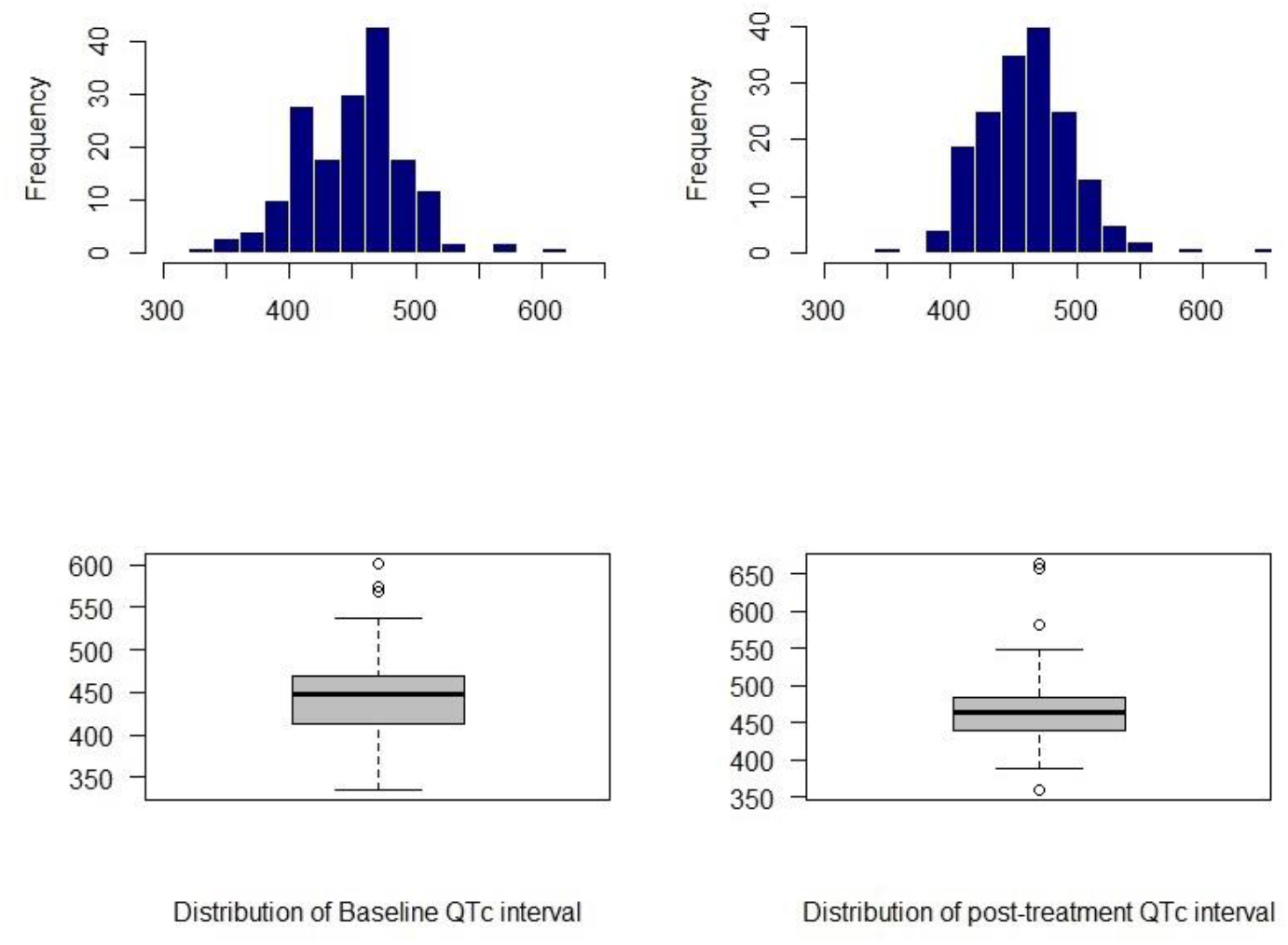
Baseline and post-treatment QTc interval in monotherapy and combination therapy group. Hydroxychloroquine alone group mean ± SD values: Baseline QTc interval = 459.9 ± 38.3, post-treatment QTc interval = 462.5 ± 43.1, ΔQTc = 2.6 ± 29.1. Hydroxychloroquine and azithromycin group mean ± SD values: Baseline QTc interval = 447.1 ± 44.1, post-treatment QTc interval = 464.3 ± 59.1, ΔQTc = 17.2 ± 53.4.

Numbers of post-treatment prolonged QTc interval (ΔQTc > 60 milliseconds or post-treatment QTc interval ≥ 500 milliseconds) were more in combination therapy group but it had no significant statistical difference than another group (35 (24.5%) vs 3 (10.3%); *P* = 0.094). Generally, 22.1% of patients had a prolonged QTc interval after treatment (Table 2).

9 patients in the population developed an abnormal heart rate after treatment (7 tachycardia and 2 bradycardia). Other events such as atrial fibrillation (3 patients), premature atrial contraction (1 patient), ST-segment elevation (4 patients), and bundle branch blocks (1 patient with right bundle branch block and 3 patients with left bundle branch block) occurred with low incidence and there was no significant statistical difference between two groups (Table 3).

**Table 3.**
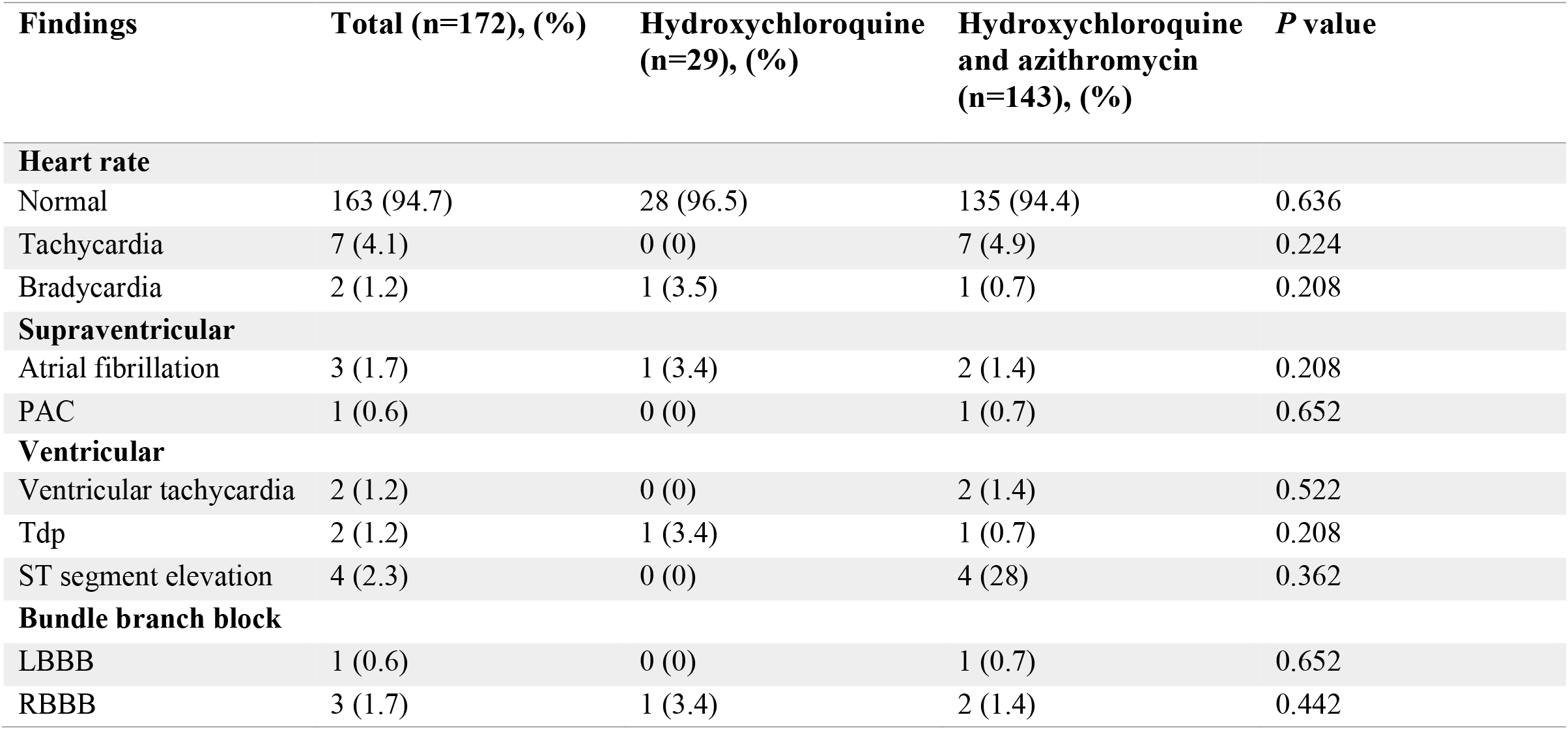
Post-treatment electrocardiographic findings of COVID-19 patients. Abbreviations: PAC, Premature atrial contraction; TdP, Torsade de pointes; RBBB, Right bundle branch block; LBBB, Left bundle branch block.

| Findings | Total (n=172), (%) | Hydroxychloroquine (n=29), (%) | Hydroxychloroquine and azithromycin (n=143), (%) | P value |
| --- | --- | --- | --- | --- |
| **Heart rate** |  |  |  |  |
| Normal | 163 (94.7) | 28 (96.5) | 135 (94.4) | 0.636 |
| Tachycardia | 7 (4.1) | 0 (0) | 7 (4.9) | 0.224 |
| Bradycardia | 2 (1.2) | 1 (3.5) | 1 (0.7) | 0.208 |
| **Supraventricular** |  |  |  |  |
| Atrial fibrillation | 3 (1.7) | 1 (3.4) | 2 (1.4) | 0.208 |
| PAC | 1 (0.6) | 0 (0) | 1 (0.7) | 0.652 |
| **Ventricular** |  |  |  |  |
| Ventricular tachycardia | 2 (1.2) | 0 (0) | 2 (1.4) | 0.522 |
| Tdp | 2 (1.2) | 1 (3.4) | 1 (0.7) | 0.208 |
| ST segment elevation | 4 (2.3) | 0 (0) | 4 (2.8) | 0.362 |
| **Bundle branch block** |  |  |  |  |
| LBBB | 1 (0.6) | 0 (0) | 1 (0.7) | 0.652 |
| RBBB | 3 (1.7) | 1 (3.4) | 2 (1.4) | 0.442 |

2 patients developed ventricular tachycardia and both of them were in a combination therapy group. 2 cases of torsade de pointes occurred (1 in the monotherapy group and 1 in the combination therapy group) (Table 3).

Male gender (27 of 87 patients [31.1%] vs 11 of 85 patients [12.9%]; *P* = 0.005), or baseline QTc ≥ 450 milliseconds (23 of 79 patients [29.1%] vs 15 of 93 patients [16.1%]; *P* = 0.43), or high-risk Tisdale score (7 of 17 patients [41.2%] vs 31 of 155 patients [20%]; P < 0.001) increased likelihood of prolonged QTc interval (QTc ≥ 500 millisecond or ΔQTc > 60 millisecond). Also, in the adjusted model of the three above risk factors, male gender and high-risk Tisdale score remained independently associated with the occurrence of prolonged QTc interval. Underlying diseases and QTc prolonging agents’ consumption did not correlate with prolonged QTc interval (Table 4).

**Table 4.** Prolonged QTc interval (corrected QT interval) risk assessment. Abbreviation: ms, milliseconds; QTc, Corrected QT interval; CI, Confidence interval; NT, Not tested.

| Characteristic | Patients numbers, (%) | Prolonged QTc interval |  |  |  |
| --- | --- | --- | --- | --- | --- |
|  |  | Odds ratio (95% CI) | P value | Adjusted odds ratio (95% CI) | P value |
| Male | 85 (49.4) | 3.027 (1.389 – 6.600) | 0.005 | 4.113 (1.772 – 9.545) | 0.001 |
| Baseline QTc ≥ 450 ms | 79 (45.9) | 2.136 (1.024 – 4.456) | 0.043 | 1.681 (0.749 – 3.774) | 0.208 |
| Congestive heart failure | 48 (27.9) | 1.068 (0.481 – 2.369) | 0.871 | NT | NT |
| Hypertension | 53 (30.8) | 0.527 (0.224 – 1.244) | 0.144 | NT | NT |
| Coronary artery disease | 11 (6.3) | 2.134 (0.590 – 7.719) | 0.248 | NT | NT |
| Diabetes mellitus | 60 (34.8) | 0.827 (0.383 – 1.786) | 0.628 | NT | NT |
| Chronic obstructive pulmonary disease | 4 (2.3) | 3.667 (0.499 – 26.639) | 0.202 | NT | NT |
| Chronic kidney disease | 10 (5.8) | 1.555 (0.382 – 6.327) | 0.537 | NT | NT |
| **QT prolonging agents** |  |  |  |  |  |
| 1 QT prolonging agent | 29 (16.9) | 1 |  | NT | NT |
| ≥2 QT prolonging agent | 143 (83.1) | 2.809 (0.801 – 9.847) | 0.107 | NT | NT |
| **Tisdale score** |  |  |  |  |  |
| Low risk (< 7) | 43 (25) | 1 |  | 1 |  |
| Moderate risk (7 – 11) | 111 (64.5) | 1.793 (0.679 – 4.733) | 0.239 | 2.196 (0.764 – 6.311) | 0.144 |
| High risk (≥ 11) | 17 (9.9) | 4.317 (1.182 – 15.760) | 0.027 | 6.369 (1.467 – 27.600) | 0.013 |

141 patients discharged and 31 patients expired; it shows no significant statistical difference between the outcomes of the monotherapy group and combination therapy group (4 death and 25 discharge in monotherapy group vs 27 death and 116 discharge in combination therapy group; *P* = 0.516).

Due to QTc prolongation, 14 patients did not continue hydroxychloroquine and azithromycin after 4 days. 15 patients developed transient nausea and dizziness that may be due to the side effects of hydroxychloroquine.

## 4 Discussion

We found that QTc interval prolongation in the combination therapy group was more than the monotherapy group but it was not statistically significant and it seems that concerns of arrhythmogenic events of azithromycin should not be determinative on choosing the drug regimen. Male gender, baseline QTc interval ≥ 450 milliseconds, and high-risk Tisdale score increased risks of QTc interval prolongation. Underlying diseases such as diabetes mellitus, congestive heart failure, and hypertension did not show effects on QT interval prolongation. Few patients developed conditions such as tachycardia, bradycardia, atrial fibrillation, ST-segment elevation, and bundle branch blocks; but the study suggests that their incidence was not impressive and two groups had no significant statistical difference.

We did not find an effective role for Torsade de pointes and ventricular tachycardia as a risk factor on patients’ outcomes (in the monotherapy group and combination therapy group).

Hydroxychloroquine and azithromycin combination were common at the beginning of the pandemic: studies in vitro report hydroxychloroquine could inhibit virus fusion and entry to cell and confronts against the virus[7, 8]. But a notable concern raised with this treatment: is the short-term administration of these drugs increasing the risk of QT interval prolongation and Torsade the pointes?

Many studies introduce antimalarial drugs as QT interval prolonging agents and among them, quinidine and halofantrine are famous[20, 21]. The function and structure of hydroxychloroquine are similar to quinidine (a class IA antiarrhythmic agent). Hydroxychloroquine can block human ether-à-go-go related gene (hERG) potassium channel, prolong QT interval, and increase the risk of Torsade de pointes which could lead to sudden cardiac death[22].

Hydroxychloroquine chronic consumption and overdose may lead to QT interval prolongation and Torsade de pointes[23, 24].

Azithromycin –a widely prescribed antibiotic– caused QT prolongation in many cases[25, 26]. In 2012 Ray reported azithromycin (within 5 days) increased sudden cardiac death (its effect was small but statistically significant); after stopping the treatment this effect did not persist[15]. Increasing the intracellular sodium ion and dysregulation of intracellular calcium may be the probable mechanism[27].

Mercuro et al. found 23% of patients who received hydroxychloroquine with or without azithromycin developed QT interval prolongation and 1 patient developed Torsade de pointes[28]. Saleh et al. reported –among 201 patients treated with hydroxychloroquine and azithromycin– QT prolongation in the hydroxychloroquine and azithromycin group, was more than hydroxychloroquine alone group and none of the patients developed Torsade de pointes[29]. Rosenberg et al. showed, the most common adverse event of hydroxychloroquine alone or with azithromycin, was abnormal electrocardiogram findings, and the combination therapy group and monotherapy group had no significant difference[11]. Our results are consistent with the above findings in some aspects such as we found 22.1% of patients developed prolonged QT interval and it was the most common adverse event, even though we did not find a significant difference in the occurring QTc interval prolongation between the combination therapy group and the monotherapy group.

We found male gender as a risk factor for QT prolongation; however, a previous study, suggested the female gender as a risk factor for adverse drug reactions[30]. This variation may be caused by the effect of the novel coronavirus at heart that we did not evaluate by a control group.

QTc interval prolongation may be led to Torsade de pointes but it is not a definitive predictive factor for Torsade de pointes occurrence. Among QT prolonging agents, anti-arrhythmic drugs have a greater role in causing Torsade de pointes (1-5%) than non-cardiac drugs (0.001%)[31]. In our study, 2 patients developed Torsade de pointes, and the incidence of Torsade de pointes between the combination therapy group and monotherapy group did not differ significantly.

Our findings suggest prescription of hydroxychloroquine and azithromycin needs careful cardiac monitoring; especially in males, patients with high-risk Tisdale score, and in patients who have baseline QTc interval ≥ 450 milliseconds. This is consistent with the American College of Cardiology recommendation, for baseline risk assessment and careful QTc interval monitoring; also, NIH (National Institutes of Health) states similar concerns by limiting the high-dose prescription of these drugs to clinical trials[12, 32].

## Limitations

It seems hydroxychloroquine and azithromycin play role in prolongation of the QT interval, but our main limitation was the absence of the control group for evaluating the electrophysiologic effects of COVID-19. The numbers of patients included in this study were fewer compare to the numbers of patients treated by hydroxychloroquine with or without azithromycin, from the initial phases of the COVID-19 pandemic. We evaluate hospitalized patients and we did not follow them after discharge to find out long-term adverse drug reactions.

## 5 Conclusion

Hospitalized patients treated by hydroxychloroquine with or without azithromycin, had no significant difference in prolongation of QT interval and outcome. But numbers of patients with prolonged QT intervals in this study emphasize careful cardiac monitoring during therapy; especially in high-risk patients. Further studies and clinical trials are needed, to evaluate the effectiveness of these drugs on the outcome and possible adverse drug reactions.

## Data Availability

The data that support the findings of this study are available from the corresponding author, upon reasonable request.

## Acknowledgments

We would like to thank Dr. Reza Ghadimi for his cooperation.

## Notes

The authors declare no competing financial interest.

## Ethical Approval

This study has been approved by the ethics committee of Babol University of Medical Sciences (Babol, Iran).

